# Carotid atherosclerosis in people of European, South Asian and African Caribbean ethnicity in the Southall and Brent Revisited study (SABRE)

**DOI:** 10.1101/2022.07.15.22277676

**Authors:** Rayan Anbar, Nish Chaturvedi, Sophie V. Eastwood, Therese Tillin, Alun D. Hughes

**Affiliations:** MRC unit for Lifelong Health & Ageing, Department of Population Science & Experimental Medicine, Institute of Cardiovascular Science, University College London, London, UK; Department of Diagnostic Radiology, Faculty of Applied Medical Sciences, King Abdul-Aziz University, Jeddah, Saudi Arabia

**Keywords:** ethnicity, cardiovascular disease, atherosclerosis, carotid artery

## Abstract

**Background:** Atherosclerotic cardiovascular disease (CVD) risk differs by ethnicity. In comparison with Europeans (EA) South Asian (SA) people in UK experience higher risk of coronary heart disease and stroke, while African Caribbean people have a lower risk of coronary heart disease but a higher risk of stroke.

**Aim:** To compare carotid atherosclerosis in EA, SA and AC participants in the Southall and Brent Revisited (SABRE) study and establish if any differences were explained by established or novel CVD risk factors.

**Methods:** Cardiovascular risk factors were measured, and carotid ultrasound was performed in 985 individuals (438 EA, 325 SA, 228 AC). Carotid plaques, and intima-media thickness (cIMT) were measured. Associations of carotid atherosclerosis with ethnicity were investigated using regression analyses, with and without adjustment for potential confounders (age, sex) and mediators (education, diabetes, hypertension, total cholesterol, HDL-C, alcohol consumption, current smoking).

**Results:** Prevalence of any plaque was similar in EA and SA, and lower in AC (17%, 17%, and 6% respectively; p < 0.001 by ANOVA). Total plaque area was also similar in EA and SA but reduced in AC, but there were no major differences in the maximum height or length of plaques in people with plaques by ethnic group. These ethnic differences were unaffected by adjustment for potential confounders or mediators. After adjustment for age and sex cIMT was higher in AC but this difference was attenuated by adjustment for CVD risk factors.

**Conclusions:** Prevalence of carotid artery atherosclerotic plaques varies by ethnicity, independent of risk factors. The similarity of plaque burden in SA and EA despite established differences in CVD risk in these ethnic groups casts some doubt on the utility of carotid ultrasound as a means of assessing risk across ethnic groups.

## Introduction

Atherosclerotic cardiovascular disease (CVD) is the leading cause of mortality and morbidity worldwide.(1) There are marked differences in CVD risk in different ethnic groups, even within the same country: for example the risk of coronary heart disease (CHD) is more than 50% higher in migrants from the Indian subcontinent than in people of European origin in UK,(2) while people of African ethnicity in the UK have markedly elevated risk of stroke, but reduced risk of CHD in comparison with Europeans or migrants from the Indian subcontinent.(3, 4) While in all ethnic groups, classical risk factors (e.g. blood pressure, total cholesterol, high-density lipoprotein cholesterol, smoking, diabetes and family history)(2) predict risk of CVD,(5) and some risk factors, such as dysglycaemia and adiposity patterns differ by ethnicity,(6, 7) differences in these factors only partially explain ethnic differences in CVD risks.(4)

Detailed phenotyping of subclinical atherosclerosis may provide more insights into ethnic differences in CVD risk. Ultrasonography is a reliable and non-invasive technique that is widely used to assess atherosclerosis in the carotid artery.(8) In addition to measurement of common carotid artery intima-media thickness (cIMT) and quantification of plaques in the carotid bifurcation,(9) this method can also provide some information on plaque composition and vulnerability.(10-12)

The present investigation compares the burden of carotid atherosclerosis in a multi-ethnic cohort of people resident in UK and investigates the potential role of established and novel cardiovascular risk factors in explaining any differences observed between ethnic groups. Individuals studied were participants in the third follow-up visit of the South and Brent Revisited (SABRE) study, a longitudinal cohort that has been followed up for over 30 years.

## Methods

### Participants and study design

SABRE is a multi□ethnic cohort of predominantly European (EA), South Asian (SA) and African Caribbean (AC) participants living in West and North London, which was established in 1988 to examine ethnic differences in chronic disease and cardiometabolic disease in particular.(13) Participants’ ethnicity was determined by interviewers based on grand-parental origin and confirmed by participants. The analysis presented here is based on only people of EA, SA and AC ethnicity. Ethical approval was obtained from Ealing, Hounslow and Spelthorne, Parkside, and University College London Research Ethics Committees and all participants provided written informed consent.

### Participant information and clinical investigations

At visit 3(2014-2018), participants were invited to a clinic appointment, which involved completing a health and lifestyle questionnaire in addition to clinic measurements.**(14)** All participants were asked to fast and refrain from alcohol, smoking, and caffeine for ≥12 h before attendance and not to take their medication on the morning of the clinic visit. Information was recorded on age, sex, health behaviours, medical history, and medication. Height and weight were measured using a standardized protocol and body composition was measured using a Tanita BC 418 body composition analyzer. Seated brachial blood pressure (BP) was measured using an appropriately sized cuff using an automatic Omron 705IT after 5-10 minutes rest according to ESH guidelines.(15) The average of the second and third recordings was used as the estimate of clinic BP. Diabetes mellitus was defined according to the 1999 WHO guidelines,(16) or physician diagnosis or receipt of anti-diabetes medications. Hypertension was defined as physician-diagnosed hypertension or participant-reported hypertension or receipt of BP-lowering medication. Smoking was classified into current or not. Alcohol consumption was categorised according to UK guidelines into none, ≤ 14 units per week or > 14 units per week. Blood and urine samples were taken, and whole blood, serum, EDTA plasma and urine stored at -80°C prior to analysis. Glycosylated haemoglobin (HbA1c) was measured on an automated platform (c311, Roche Diagnostics, Burgess Hill UK), serum total cholesterol (TC) and high-density lipoprotein cholesterol (HDL-C) and triglycerides were using enzymatic methods (Roche/Hitachi cobas c system). Low density lipoprotein cholesterol (LDL-C), Medium high density lipoprotein (HDL), apoB/apoAI ratio, albumin docosahexaenoic acid, histidine, tyrosine, glutamine were measured using a high-throughput nuclear magnetic resonance (NMR) platform.(17) Urinary albumin:creatinine ratio was measured with an immunoturbidimetric assay combined with a kinetic colorimetric assay (Roche/Hitachi cobas c system) and NT-pro BNP, C-reactive protein (CRP), and high sensitivity troponin T (TnT) were measured on an automated platform (e411, Roche Diagnostics, Burgess Hill UK). IL-6 was measured using a high sensitivity ELISA assay (R&D Systems, Biotechne, Oxon, UK). All assays used the manufacturers calibration and quality control material.

### Ultrasound Measurements

Ultrasound scans were performed by an experienced sonographer using a GE Vivid I Ultrasound system equipped with a 6-13MHz broadband linear array transducer (12L-RS). The common carotid artery (CCA), internal carotid artery (ICA), external carotid artery (ECA) was assessed along the long- and short-axes bilaterally. 2D-images, spectral-Doppler imaging, colour and power doppler were recorded. Adequate quality of ECG signals and ultrasound images was ensured throughout the examination. A cine loop of at least five cardiac cycles at three angles (lateral, posterior, and anterior) as well as one 8-bit greyscale image captured at the R wave for each angle were acquired. cIMT and carotid lumen diameter was measured from the best visualised image over a 10mm segment in the common carotid artery according to the American Society of Echocardiography Carotid Intima-Media Thickness Task Force Consensus Statement.(18) Plaque was defined according to the Mannheim consensus(19, 20) as a focal lesion that encroached into the carotid artery lumen by ≥0.5 mm or ≥50% of the surrounding cIMT value or had a thickness >1.5 mm as measured from the media-adventitia interface to the intima-lumen boundary. Carotid stenosis >50% was assessed by visual inspection of the ultrasound scan using color and power Doppler imaging as needed and quantified according to NASCET criteria.(21) All analyses were performed offline using validated software (AMS II)(22) that included automated measurement of plaque characteristics and estimation of grey-scale median (GSM).(23, 24) Repeatability and reproducibility of cIMT and plaque characteristics have been reported previously.(14)

### Statistical analysis

Statistical analyses were performed with Stata v.17.1 (StataCorp, College Station, TX, USA). Continuous data for the sample were summarised as means and standard deviations (SD) or median (interquartile range) for skewed data, categorical data as counts and percentages. Normality was assessed through frequency histograms, Q plots and Shapiro-Wilk tests. Comparisons between ethnic groups were made using multivariable regression modelling. Four models were used: Model 1) unadjusted; Model 2) age and sex; Model 3) Model 2 plus adjusted for established potential mediators (education, diabetes, hypertension, total cholesterol, HDL-C, alcohol consumption, current smoking); Model 4) Model 3 minus total cholesterol and HDL-C plus novel potential mediators (LDL cholesterol, medium HDL, apoB/apoAI ratio, albumin docosahexaenoic acid, histidine, tyrosine, glutamine, urinary albumin creatinine ratio, NT-pro BNP, CRP, TnT). Supplementary models were also examined where BMI or waist hip ratio were added, or diabetes was replaced by HbA1c, CRP was replaced by IL6, and hypertension by systolic blood pressure. Choice of covariates was based on a priori knowledge(25-28).

The possibility of effect modification by sex was looked for in all models by including a sex x ethnicity interaction term, if this was not statistically significant both sexes were pooled for analysis, otherwise it was planned that results for both sexes would be analysed separately. Dichotomous variables (e.g. presence of plaque or presence of carotid stenosis >50%) were modelling using multivariable logistic regression, and ordinal logistic regression for ordered categorical variables with fewer than six categories (median plaque grade (manual and automatic)); odds ratios (OR) or marginal probabilities and 95% confidence intervals (CI) were estimated from these models. Multiple linear regression models were used for continuous plaque measures (maximum plaque height, maximum plaque length, number of plaques, total area of plaques, lowest GSM of all plaques, cIMT) and marginal means and CI estimated. Assumptions of linearity were checked by examination of residuals and if necessary it was planned that nonlinear models would be constructed using fractional polynomials.

Inference was based on a combination of p-values, effect sizes and CI, no adjustment was made for multiple comparisons.

## Results

Table 1 shows the basic characteristics of the sample stratified by ethnicity. Participants were aged between 40 and 69 years and comprised 437 EA (mean age 74 years, 62% male), 326 SA (mean age 73.2 ± 6.3 years, 59.3% male) and 228 AC (mean age 71 years, 35.6% male). On average SA were slightly younger than EA and AC were younger than both EA and SA, and there were more women in the AC sample. AC and SA people were shorter, had higher systolic BP and more diabetes and hypertension than EA. Compared with EA, SA had a higher prevalence of known CHD, greater number of years of education, lower heart rate, lower fat %, and were less likely to be current smokers, and less likely to consume high quantities of alcohol, while AC had a lower prevalence of CHD, had a higher fat %, higher diastolic BP, more diabetes and hypertension and were less likely to consume high quantities of alcohol.

**Table 1.**
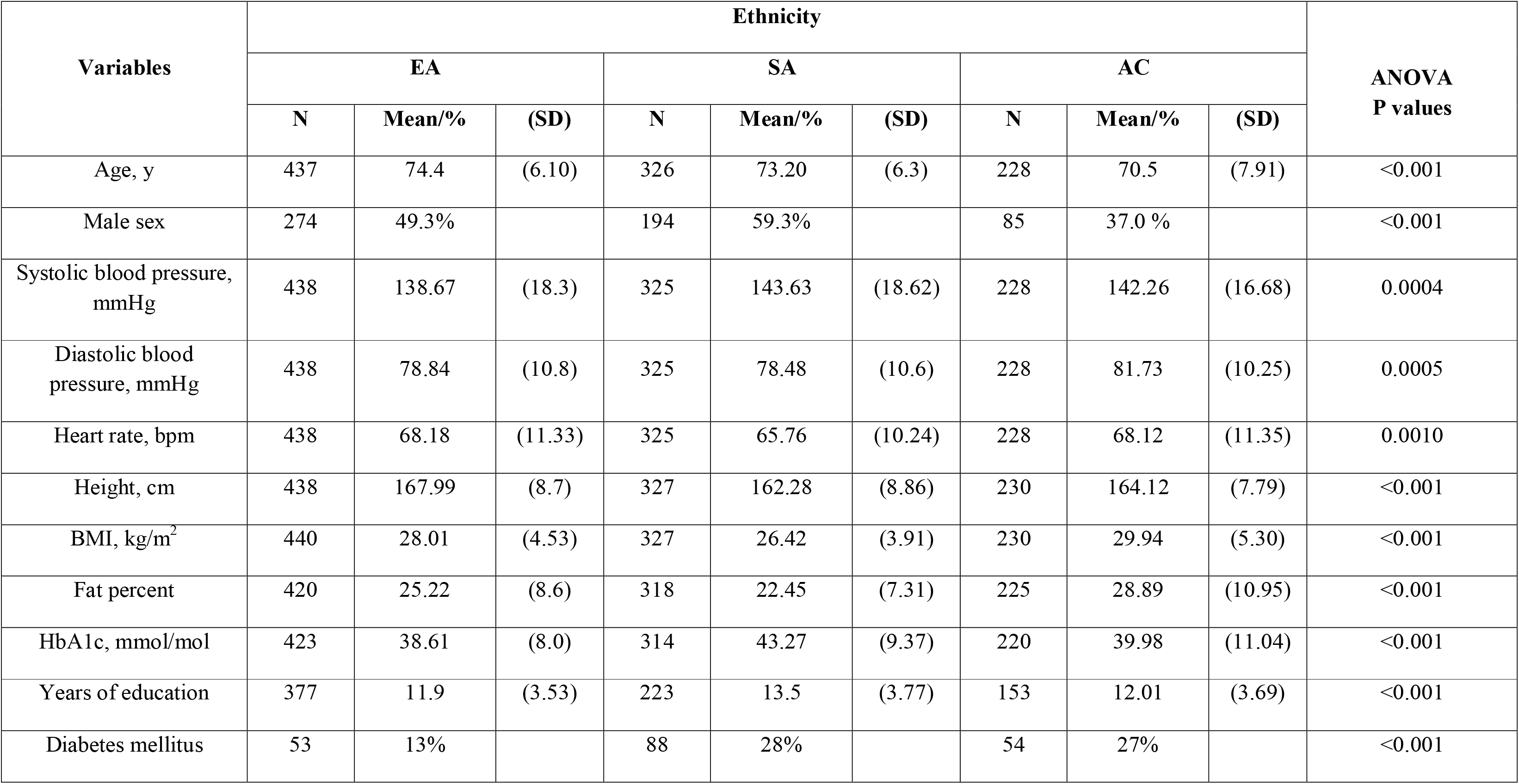
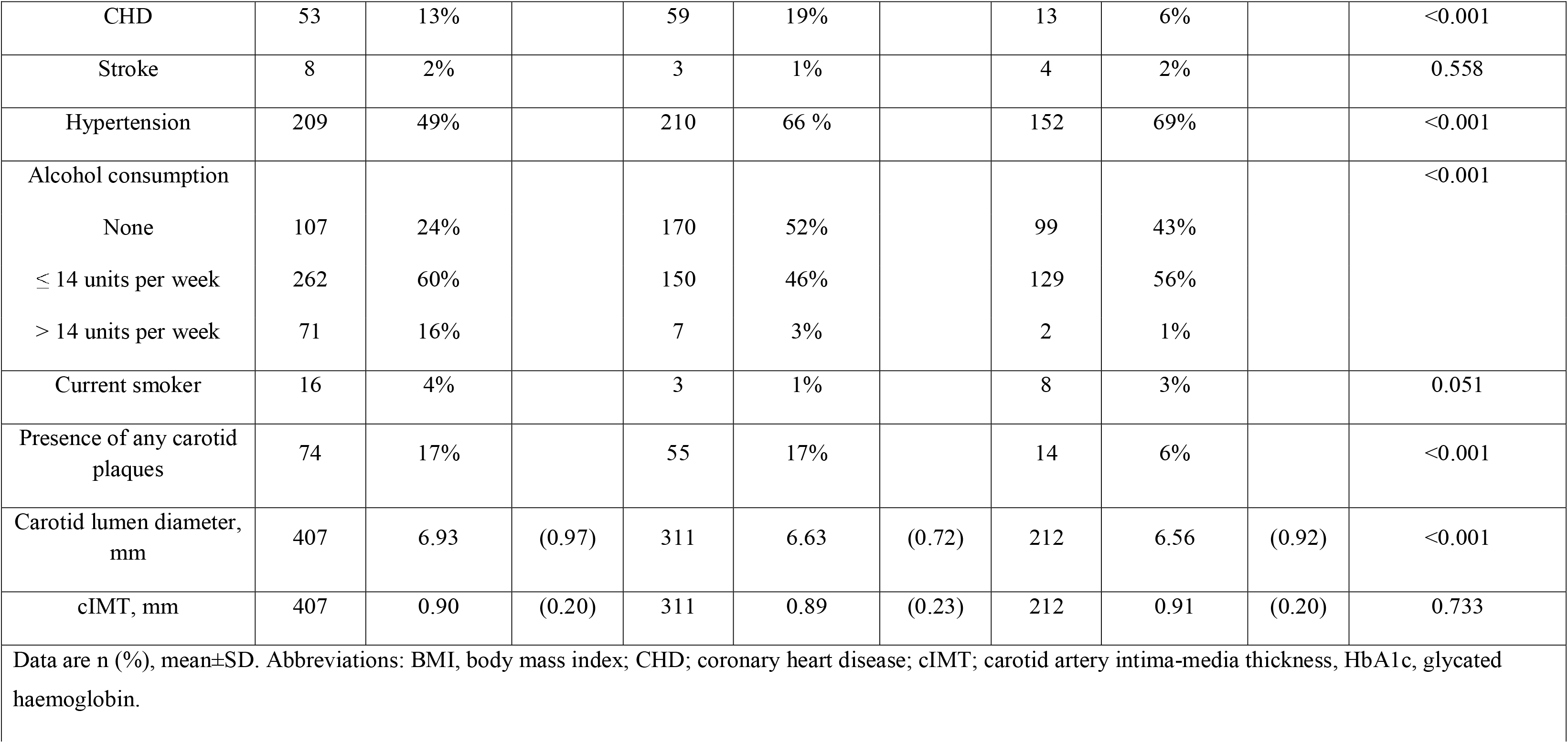
Characteristics of sample by ethnicity

cIMT was similar by ethnicity in an unadjusted model (**Table 1**), but plaques were more frequent in EA and SA than AC; however there was no difference between EA and SA (17% of EA, 17% of SA and 6% of AC had at least one carotid plaque; **Table 1**). Plaques were more common in men than women but there was no evidence that sex modified the ethnic difference in plaque prevalence (**Figure 2**). There were no marked differences in distribution of plaques by ethnicity: 57.5% of Europeans had plaques in the left carotid artery, 47.6% in the right carotid artery and 44.1% had plaques bilaterally. 42.5% of South Asians had plaques in the left carotid artery, 44.1% in the right carotid artery and 32.7 % had plaques bilaterally. 10% of African Caribbean’s had plaques in left of carotid artery, 8.1% in the right carotid artery and 23.0 % had plaques bilaterally (**Figure 1**). The marginal probabilities of having one or more plaques in each ethnic group are shown in (**Table 2)** with and without adjustment for potential confounders or mediators. The probability of having one or more plaques was similar in EA and SA but was lower by around 50% in AC, statistical adjustment had little effect on these estimates.

**Figure 1.**
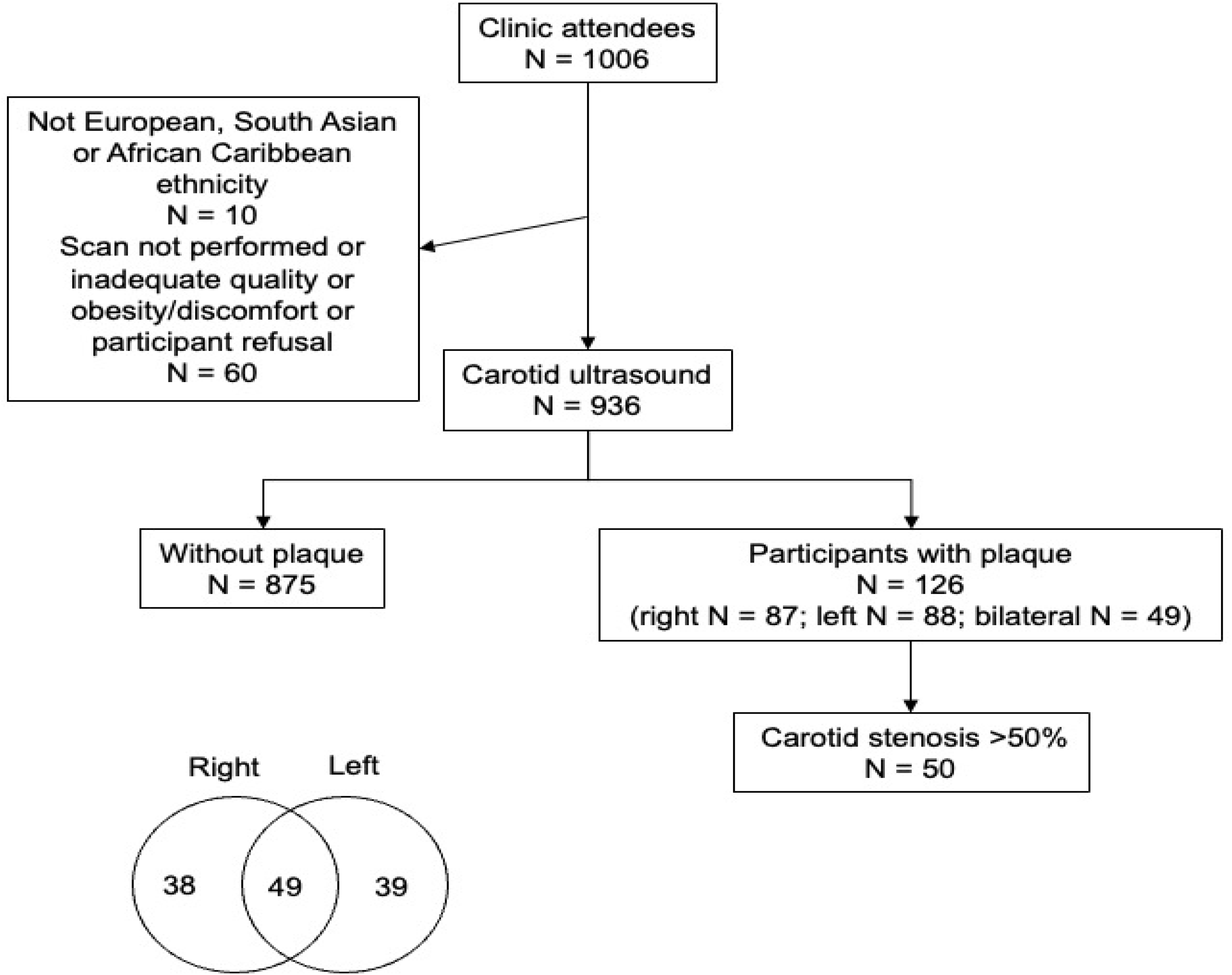
Diagram of participant flow including location of plaques as a Venn diagram.

**Figure 2.**
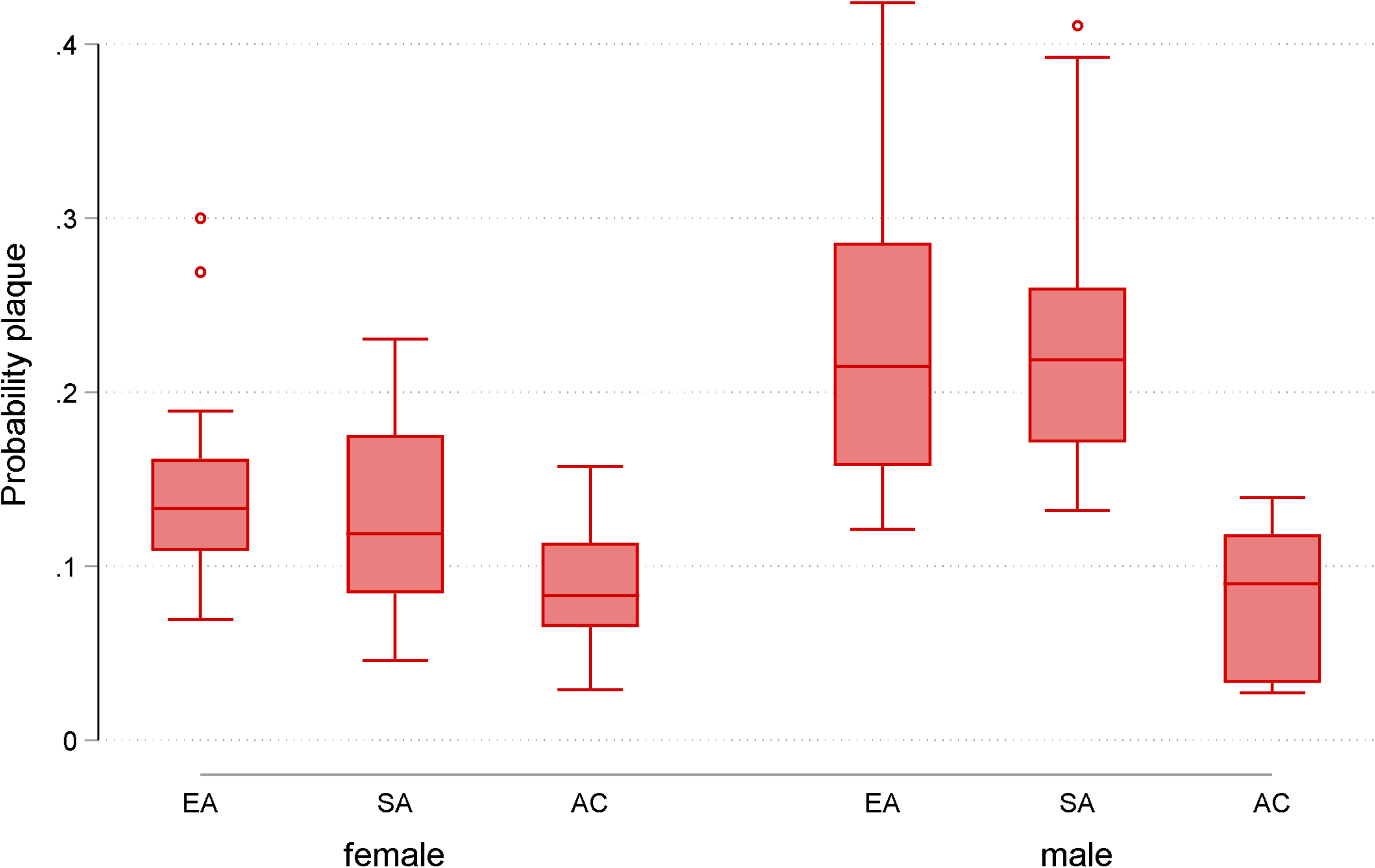
Boxplot showing probability (adjusted for age) of having a plaque by sex and ethnicity.

**Table 2.** Marginal probabilities of having any plaques by ethnicity with and without adjustment for potential confounders or mediators

| Prevalence of plaque (95% CI) by ethnicity |  |  | P-value |  |  |
| --- | --- | --- | --- | --- | --- |
| EA | SA | AC | EA v SA | EA vs AC | SA vs AC |
| Model 1 |  |  |  |  |  |
| 0.16<br>(0.13, 0.20) | 0.16<br>(0.12, 0.20) | 0.06<br>(0.03, 0.09) | 0.992 | <0.001 | <0.001 |
| Model 2 |  |  |  |  |  |
| 0.15<br>(0.12, 0.18) | 0.16<br>(0.12, 0.20) | 0.07<br>(0.03, 0.11) | 0.626 | 0.008 | 0.004 |
| Model 3 |  |  |  |  |  |
| 0.16<br>(0.12, 0.19) | 0.16<br>(0.12, 0.20) | 0.08<br>(0.04, 0.13) | 0.972 | 0.025 | 0.005 |
| Model 4 |  |  |  |  |  |
| 0.17<br>(0.13, 0.21) | 0.16<br>(0.12, 0.20) | 0.07<br>(0.03, 0.11) | 0.75 | 0.003 | 0.012 |

Model 1: unadjusted, Model 2 age, sex adjusted, Model 3: Model 2 + education, diabetes, total cholesterol, HDL, alcohol consumption, current smoking, hypertension, statin use, Model 4: model 3+ NT pro BNP, C-reactive protein, interleukin-6, troponin, low density cholesterol, medium high-density lipoprotein, ApoB/ApoAI ratio, albumin, histidine, tyrosine, glutamine, docosahexaenoic acid, urinary albumin creatinine ratio, estimated glomerular filtration rate.

After adjustment for age and sex cIMT was higher in AC than EA or SA but there was no difference in cIMT between EA and SA (**Table 3B**). Further adjustment for established and novel risk factors attenuated but did not abolish differences by ethnicity (**Table 3C & 3D**).

**Table 3A.**
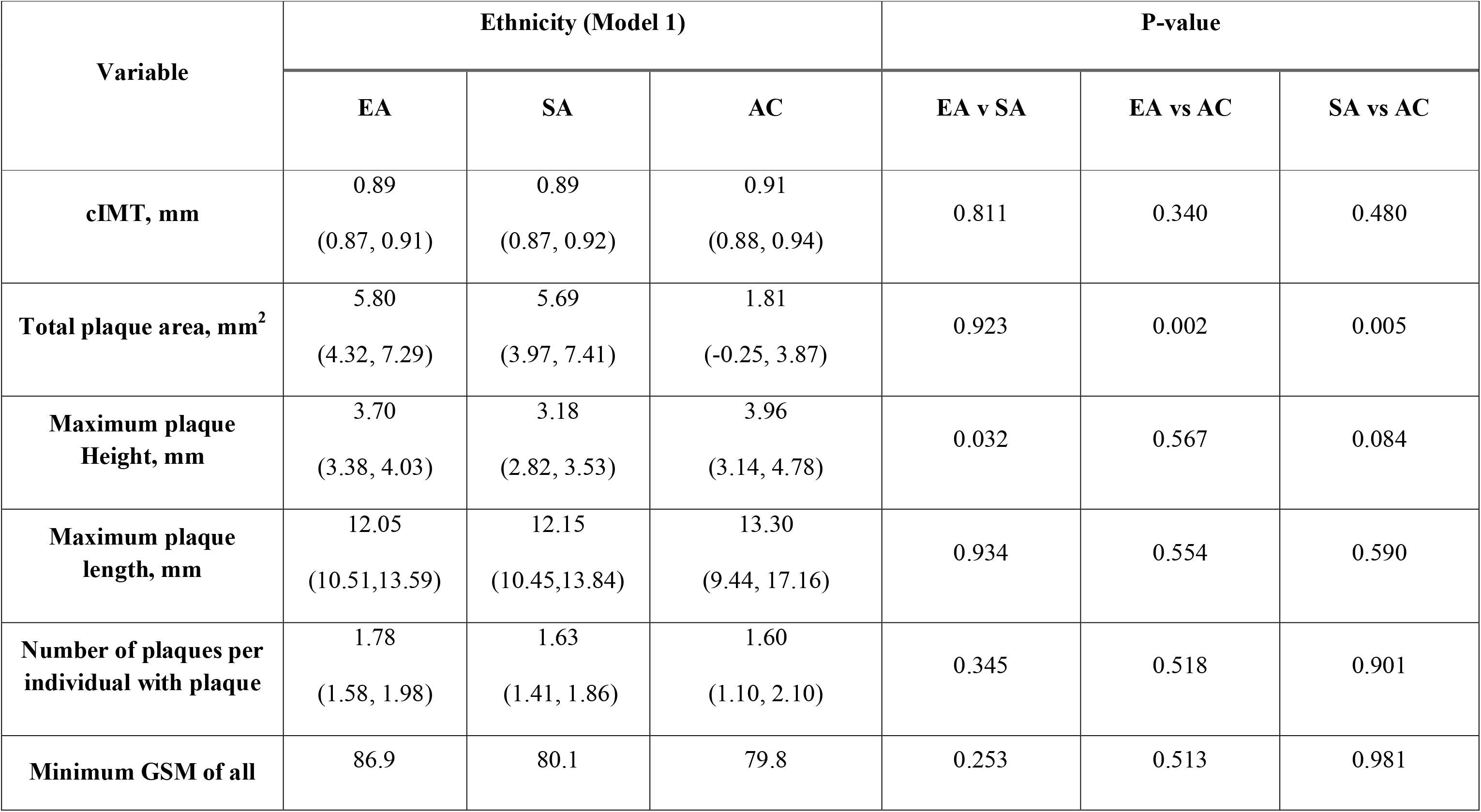
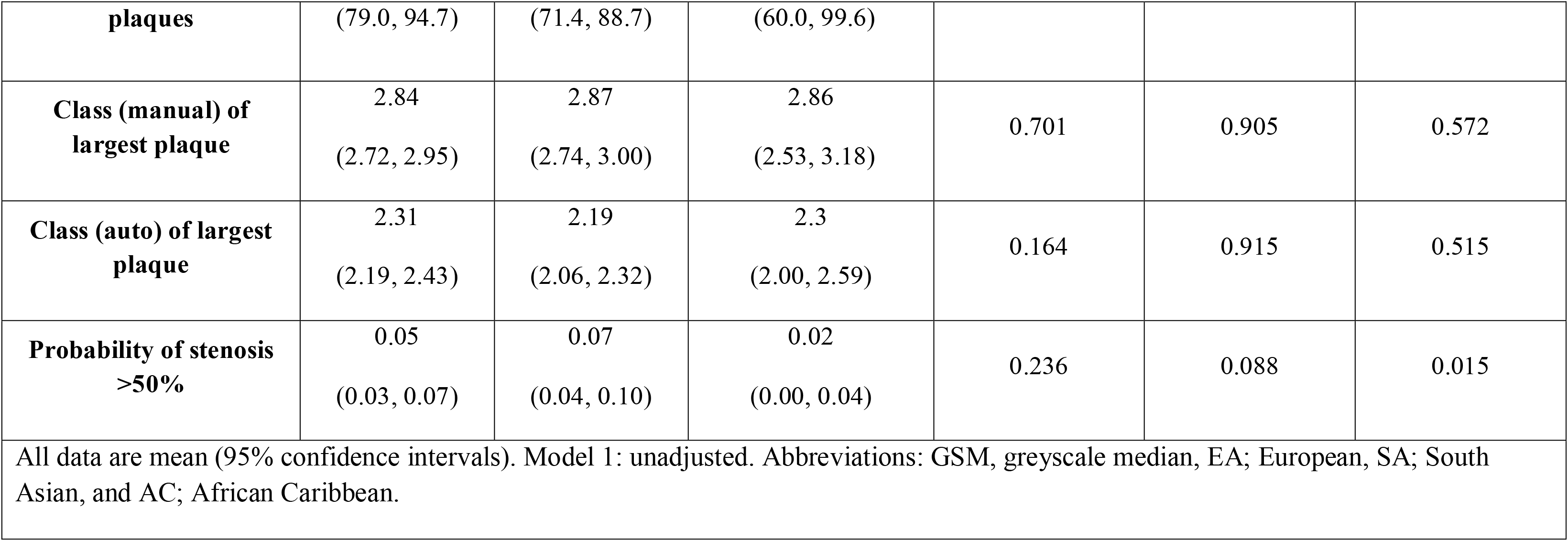
cIMT and plaque characteristics by ethnicity without adjustment (Model 1)

**Table 3B.**
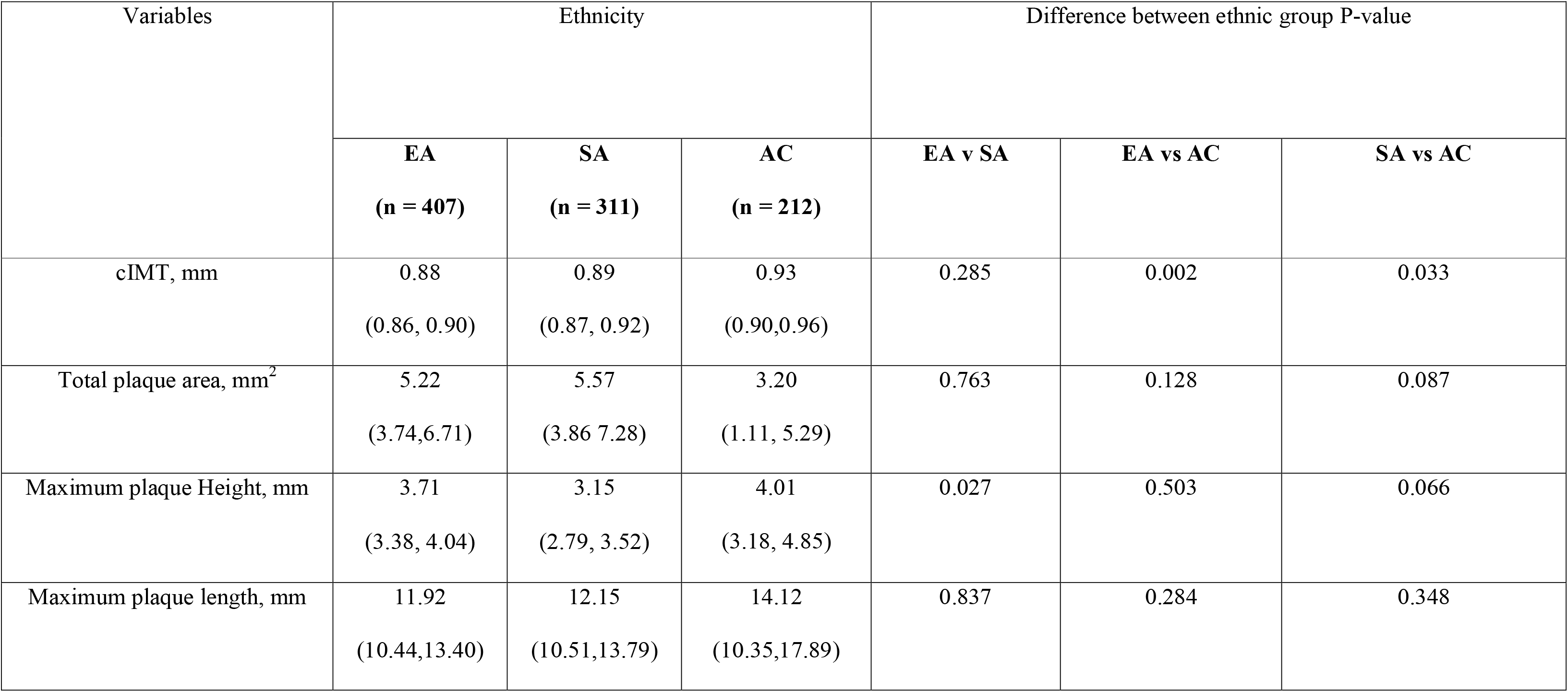
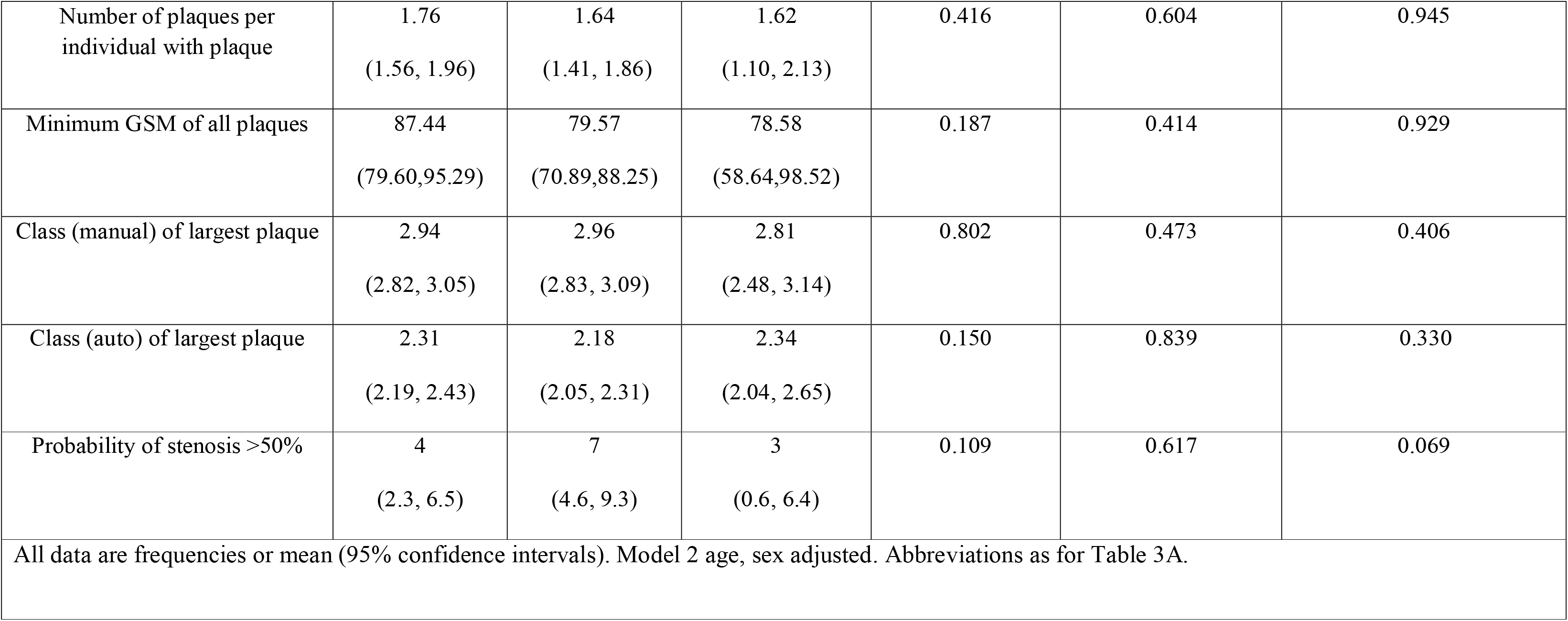
Marginal means of plaque characteristics by ethnicity with adjustment for age and sex (Model 2)

**Table 3C.**
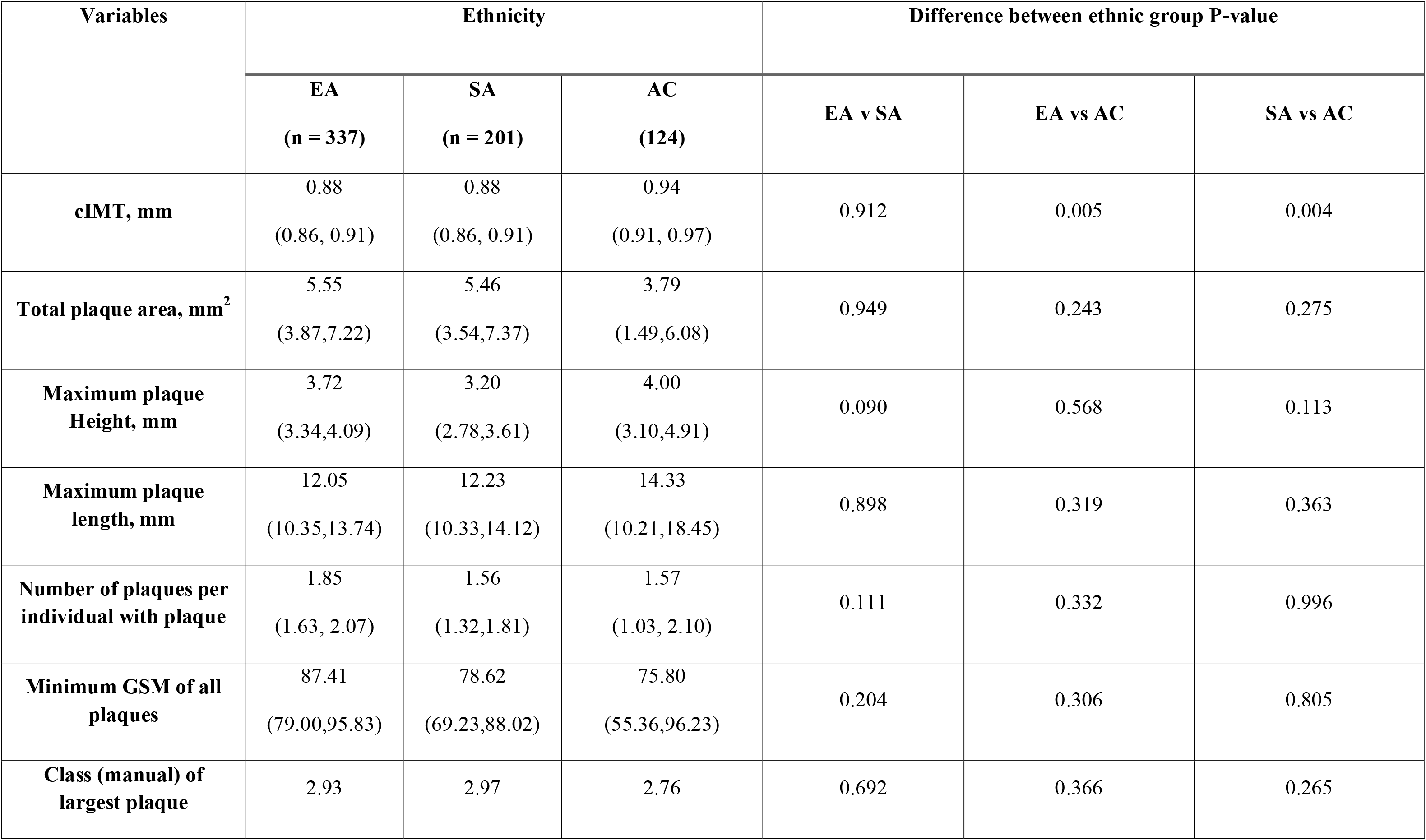
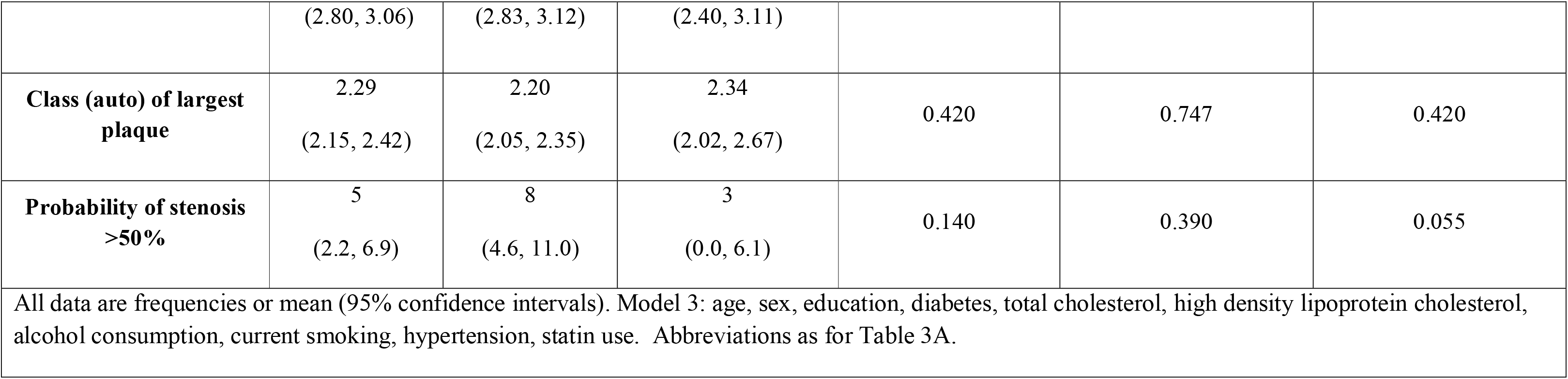
Marginal means of carotid intima-media and plaque characteristics by ethnicity after adjustment for potential confounders and mediators (Model 3)

**Table 3D.** cIMT and plaque characteristics by ethnicity after adjustment for novel risk factors (Model 4).

| Variables | Ethnicity |  |  | P-value |  |  |
| --- | --- | --- | --- | --- | --- | --- |
|  | EA<br>(n = 330) | SA<br>(n = 195) | AC<br>(n = 122) | EA v SA | EA vs AC | SA vs AC |
| cIMT, mm | 0.89<br>(0.86, 0.91) | 0.89<br>(0.86, 0.91) | 0.93<br>(0.90, 0.96) | 0.943 | 0.043 | 0.058 |
| Total plaque area, mm<sup>2</sup> | 5.34<br>(4.15,7.23) | 5.55<br>(3.52,7.33) | 3.73<br>(0.32, 5.1) | 0.881 | 0.318 | 0.304 |
| Maximum plaque Height, mm | 3.67<br>( 3.35,4.00) | 3.28<br>(2.75,3.52) | 3.91<br>(3.15,4.96) | 0.285 | 0.594 | 0.295 |
| Maximum plaque length, mm | 12.2<br>( 10.61,13.95) | 12.1<br>(10.2,14.1) | 13.1<br>(8.56, 17.7) | 0.821 | 0.636 | 0.758 |
| Average number of plaques per individual with plaque | 1.80<br>( 1.59, 2.01) | 1.55<br>( 1.30, 1.79) | 1.53<br>( 1.15, 2.31) | 0.216 | 0.404 | 0.922 |
| Minimum GSM of all plaques | 87.12<br>(78.12, 94.12) | 78.50<br>(69.12,87.87) | 77.53<br>(57.37,101.6) | 0.242 | 0.448 | 0.948 |
| Class (manual) of largest plaque | 2.80 | 2.94 | 2.70 | 0.269 | 0.344 | 0.669 |
|  | (2.82, 3.07) | (2.80, 3.09) | (2.46, 3.24) |  |  |  |
| **Class (auto) of largest plaque** | 2.34<br>(2.22, 2.46) | 2.19<br>(2.05, 2.33) | 2.20<br>(1.87, 2.53) | 0.762 | 0.889 | 0.740 |
| **Probability of stenosis >50%** | 5<br>(2.7, 7.0) | 6<br>(3.8, 9.2) | 3<br>(0.1, 6.9) | 0.275 | 0.523 | 0.673 |
| All data are frequencies or mean (95% confidence intervals). Model 4: model 3 + NT pro BNP, CRP, IL6, troponin-T, LDL cholesterol (directly measured) instead of total cholesterol, total cholesterol in medium HDL instead of HDL-cholesterol, ApoB/ApoAI ratio, albumin, histidine, tyrosine, glutamine, docosahexaenoic acid, urinary albumin creatinine ratio, eGFR. Abbreviations as for Table 3A. |  |  |  |  |  |  |

A comparison of plaque area, maximum plaque height, average number of plaques, minimum greyscale median, and the manual and automatic plaque class is shown in **Tables 3(A-D)**. Total plaque area was similar in EA and SA but considerably lower in AC, adjustment for age and sex, slightly attenuated the difference between EA or SA and AC, but a substantial difference remained, and this was essentially unaltered by further adjustment for potential mediators (Table 3 B-D). Echogenicity as assessed by GSM was similar in EA and SA and lower in AC. Maximum plaque height in those with plaque showed no difference among three different ethnic group; however, the maximum plaque length in AC was longer compared to EA & SA groups, despite AC having the lowest frequency of plaques. There were no differences between ethnic groups for plaque grade either using an automated or manual method. The proportions of carotid stenosis >50% were slightly higher in SA (7%) compared to EA (5%) and AC (2%), although due to the low prevalence of carotid stenosis any differences could have arisen by chance.

## Discussion

We found ethnic differences in the burden of carotid plaque and cIMT in a population-based sample of people in UK. People of AC ethnicity had a lower occurrence of carotid plaque and a smaller total plaque area than the other ethnic groups while the burden of plaque in SA and EA was similar. In those with plaque, plaque characteristics differed little between ethnic groups. Ethnic differences in plaque were unexplained by disparities in conventional and novel CVD risk factors between different ethnic groups. It is therefore unclear why the AC group had a lower prevalence of carotid plaques than the other ethnic groups. However, this observation is consistent with previous work, including in SABRE, showing lower risk of CHD in people of AC ethnicity in UK this was not explained by conventional CVD risk factors. (29, 30) cIMT was higher in AC compared with the other ethnic groups, which could be consistent with their higher risk of stroke but inconsistent with their lower risk of CHD; this difference was attenuated after adjustment for CVD risk factors.

Better understanding and assessment of the prevalence of atherosclerosis and its relationship to cardiovascular risk factors in different ethnic groups is important. Such relationships may also provide insights into the pathogenesis of atherosclerosis in all ethnic groups. Our failure to identify factors explaining ethnic differences in carotid atherosclerotic plaque despite extensive adjustment for conventional and novel risk factors suggests that important determinants of atherosclerosis remain to be identified. Mechanisms related to population migration,(31) socio-economic disadvantage(32) and racism(32, 33) seem plausible explanations, but given the differences observed between minority ethnic groups in this study this question merits further study. We cannot exclude genetic differences between populations of difference ancestry but currently there is little or no evidence to suggest that genetics makes an important contribution to ethnic differences in susceptibility to CVD. (34, 35)

Previous studies have examined ethnic differences in carotid atherosclerosis, although few have included SA people. A UK community-based study found higher cIMT and lower prevalence of plaque in AC compared with EA (36) and this difference remained after adjustment for conventional CVD risk factors. Another UK-based study observed marginally higher cIMT in EA compared with SA despite higher prevalence of CVD in SA. (37) In the US, the Multi-Ethnic Study of Atherosclerosis found that cIMT was higher in people of African American ethnicity, but the risk of new plaque formation was lower in African American, Hispanic and Chinese ethnicities compared with White Americans after adjustment for traditional CVD risk factors.(38) The Diabetes heart study also found that African American people with T2DM had higher cIMT but lower prevalence of carotid plaque compared with those of European ancestry.(39) In contrast, the Northern Manhattan Stroke study found similar maximum internal carotid artery plaque thickness (MICPT) in stroke-free African- and European-ethnicity individuals but lower MICPT in people of Hispanic ethnicity. (40) Overall, despite some inconsistencies the results of these previous studies appear in keeping with our findings.

As has been observed in some previous studies, (36-38) cIMT corresponded poorly with known risks of CVD, especially CHD, in the ethnic groups. Plaque prevalence was consistent with the known lower risk of CHD in AC, but not with the elevated risk of CVD in SA or the elevated risk of stroke in AC. (41, 42) This raises questions about the reliability of cIMT and plaque as a screening tool for early detection of atherosclerosis *across* different ethnic groups. For cIMT it has previously been suggested that arterial wall remodelling in response to hemodynamic stresses might complicate interpretation (43), but it is not obvious that this could explain the ethnic discordance between CVD risk and plaque prevalence, given the latter is generally considered a better predictor of CVD risk .(44)

This study has limitations and strengths: it is cross-sectional so causal conclusions cannot be made. Participants were drawn from a randomly selected population-based cohort but possible bias due to non-participation, attrition, missing data and residual confounding by unmeasured or imprecisely measured variables cannot be excluded. Our categorization of ethnicity is crude and may obscure important differences within ethnic categories,(45) but our categories reflect the original study design and correspond to the broad ethnic groups in used by the UK classification scheme.(46) AC participants mostly migrated between 1950 and 1960 (i.e. around the ages of 20 to 30), while most of the SA participants arrived in the UK in the 1970’s (i.e. around 40 year old) and limited data was available about exposures, including childhood exposures and healthcare provision, that occurred prior to migration. The study’s strength is first and foremost its community-based methodology. SA and AC participants make up the majority of British first-generation migrants and unlike in some countries the availability of universal health care in UK may lessen, though not abolish disadvantages in health access.(47) All examinations were conducted according to a strict approach, resulting in a comprehensive phenotyping of this older age sample

## Conclusions

EA and SA have a higher burden of atherosclerosis plaques in carotid arteries, when compared with AC but cIMT was higher in AC than other ethnicities. These differences were unexplained by a range of conventional and novel risk factors and our observations raise questions about the reliability of carotid ultrasound as a tool to predict risk in multi-ethnic populations.

## Data Availability

SABRE data is available to approved researchers upon reasonable request, details can be found at: https://www.sabrestudy.org/home-2/datasharing/.

## Conflict of Interest Statement

The authors declare that the research was conducted in the absence of any commercial or financial relationships that could be construed as a potential conflict of interest.

All authors contributed to study design and interpretation and approved the final manuscript. Rayan Anbar had full access to all the data in the study, performed the statistical analyses, and wrote the first draft of the manuscript.

## Acknowledgements

The SABRE study was funded at baseline by the Medical Research Council, Diabetes UK, and the British Heart Foundation. At follow-up the study was funded by the Wellcome Trust (067100, 37055891, and 086676/7/08/Z), the British Heart Foundation (PG/06/145, PG/08/103/26133, PG/12/29/29497, and CS/13/1/30327), and Diabetes UK (13/0004774). AH receives support from the British Heart Foundation, the Horizon 2020 Framework Programme of the European Union, the National Institute for Health Research University College London Hospitals Biomedical Research Centre, the UK Medical Research Council, the Wellcome Trust, and works in a unit that receives support from the UK Medical Research Council. SE was funded by a Diabetes UK Sir George Alberti Research Training Fellowship (Grant No. 17/0005588). We are extremely grateful to all the people who took part in the study, and past and present members of the SABRE team who helped to collect the data.

## Funding

RA is supported by a PhD scholarship grant from King Abdul-Aziz University.

## Notes

### Competing Interest Statement

The authors have declared no competing interest.

### Author Declarations

Ethical approval was obtained from Ealing, Hounslow and Spelthorne, Parkside, and University College London Research Ethics Committees and all participants provided written informed consent.

